# Cholinergic neurotransmission in the anterior cingulate cortex is associated with cognitive performance in healthy older adults: Baseline characteristics of the Improving Neurological Health in Aging via Neuroplasticity-based Computerized Exercise (INHANCE) trial

**DOI:** 10.1101/2024.10.30.24316439

**Authors:** Ana de Figueiredo Pelegrino, Mouna Attarha, Paule-Joanne Toussaint, Lydia Ouellet, Sarah-Jane Grant, Thomas Van Vleet, Etienne de Villers-Sidani

## Abstract

Aging is associated with dysfunction in the cholinergic system, including degeneration of basal forebrain cholinergic terminals that innervate the cortex, which directly contributes to age- and disease-related cognitive decline. In this study, we used [18F]fluoroethoxybenzovesamicol ([18F]FEOBV) positron emission tomography (PET) imaging to assess the effect of age on cholinergic terminal integrity in predefined regions of interest and its relationship to cognitive performance in healthy older adults who underwent neuropsychological assessment and FEOBV PET brain imaging. Our results showed age-related reductions in FEOBV binding, particularly in the anterior cingulate cortex-our primary region of interest-as well as in the striatum, posterior cingulate cortex, and primary auditory cortex. Notably, FEOBV binding in the anterior cingulate cortex was positively correlated with cognitive performance on the NIH EXAMINER Executive Composite Score. These findings suggest that [18F]FEOBV PET imaging can be used as a reliable biomarker to assess cholinergic changes in the human brain and indicate that preserving the cholinergic integrity of the basal forebrain may help maintain cognitive function and protect against age-related cognitive decline.

## 1. INTRODUCTION

The process of natural aging is associated with moderate structural and functional degeneration in the basal forebrain cholinergic neurotransmitter system.^1,2^ These age-related changes in cholinergic neurotransmission are thought to contribute to the decline in cognitive functions that are commonly observed with aging.^3–6^ Indeed, cholinergic projections from the basal forebrain play a crucial role in cognitive performance due to their involvement in high-order cognitive functions such as attention, learning and memory, and executive function.^3,6,7^ For example, enhancing cholinergic signaling has been demonstrated to enhance attention and memory performance in individuals with naturally lower cognitive performance.^8–10^ Consequently, cholinergic signaling has been identified as a potential key factor in age-related cognitive diseases, such as mild cognitive impairment (MCI) and Alzheimer’s disease.^3,11^ For instance, degeneration of basal forebrain cholinergic neurons can occur years before the onset of cognitive symptoms and may predict both cortical pathology and memory impairment.^12,13^ Understanding age-related changes in the cholinergic system is crucial for elucidating the contribution of aging to cognitive decline and for identifying those who are at risk for age-related cognitive diseases marked by cholinergic degeneration.

[18F]Fluoroethoxybenzovesamicol ([18F]FEOBV) is a reliable positron emission tomography (PET) tracer for the direct assessment of cerebral cholinergic neurotransmission in vivo. This high-affinity tracer binds to the vesicular acetylcholine transporter (VAChT), thereby enabling precise measurement of presynaptic cholinergic terminal density. ^14^ The binding pattern of [18F]FEOBV across cortical and subcortical areas reflects the unique organization of the cholinergic system, with decreases in binding indicating regions that show reduced cholinergic terminal densities due to natural aging ^15^ and age-related cognitive diseases. ^16–18^ For example, substantial declines in FEOBV binding have been documented across the entire cortex in individuals with Alzheimer’s disease, with higher binding levels associated with superior performance on global cognitive assessments.^19,20^ Similarly, reductions in cortical FEOBV binding have been observed in individuals with MCI, with binding levels positively correlating with executive function and attention in a cohort that included MCI patients and cognitively intact older adults.^18^ In natural aging, a recent study found reduced FEOBV binding in the anterior cingulate cortex of healthy older compared to younger subjects.^15^ The anterior cingulate cortex plays a key role in attention, memory, and executive function. ^21–23^ Notably, greater anterior cingulate thickness has been associated with successful cognitive aging,^24^ whereas atrophy in this region has been linked to impaired cognitive function.^25^

This paper presents the baseline characteristics of the intent-to-treat population in the Improving Neurological Health in Aging via Neuroplasticity-based Computerized Exercise (INHANCE) trial. The INHANCE trial is a Phase IIb double-blind randomized controlled study targeting community-dwelling healthy older adults aged 65 and above. Participants were randomized to receive either 35 hours of a computerized speed and attention brain training intervention (BrainHQ) or an active control involving computerized games over a 10-week period. We used [18F]FEOBV PET imaging with volume-of-interest analysis, focusing on a priori-selected regions of interest, to investigate two aspects in the largest known sample of neurocognitively intact older human brains: (1) the impact of age on cholinergic neurotransmission, and (2) the relationship between cholinergic neurotransmission and cognitive performance. Considering previous findings, we anticipated age-related reductions in FEOBV binding in the primary region of interest, the anterior cingulate cortex. Second, we anticipated that higher FEOBV binding in the anterior cingulate cortex would be associated with better cognitive performance.

## 2. METHODS

### 2.1 Study Design

Study staff supporting participants remained unblinded, while blinded staff conducted assessments, scoring, follow-ups, and baseline data analyses.

The INHANCE trial took place at McGill University, Canada, where the FEOBV radiotracer was both synthesized and administered. Below, we outline the methodological details pertinent to this paper. A comprehensive description of the training programs, measures, and data analyses can be found in the study protocol. ^26^

### 2.2 Participants

Inclusion criteria comprised individuals aged 65 or older, proficient in English or French, cognitively intact with a Montreal Cognitive Assessment (MoCA) score of ≥ 23 ^27^, and capable of fulfilling study requirements. Exclusion criteria encompassed neurocognitive disorders, suicidal ideation, major depression scoring >10 on the Geriatric Depression Scale – Short Form (GDS-SF), prior experience with BrainHQ within the past 5 years, concurrent clinical trial participation, pregnancy, substance abuse, neuroimaging contraindications, or medical conditions hindering study engagement.

The study protocol was developed in accordance with the Declaration of Helsinki guidelines and was approved by WIRB (IRB00000533) and REB of McGill University Health Centre (2020-6474). The radioligand FEOBV was approved by Health Canada (Control # 252085). All participants provided written, informed consent.

### 2.3 Imaging acquisition

All participants underwent a structural T1-weighted MRI scan (3T Siemens Prisma) and a [18F]FEOBV-PET scan using the Siemens High-Resolution Research Tomograph (HRRT; full width at half maximum of 2.4 mm) at the McConnell Brain Imaging Centre of the Montreal Neurological Institute-Hospital. The [18F]FEOBV precursor (ABX Advanced Biochemical Compounds, GmbH, Germany) was synthesized on the same day as participant testing at the cyclotron facility. Radiochemical purity was ensured using high-performance liquid chromatography. Mass dosages did not exceed the proposed limits of 0.0175 mcg/kg.^14^ Participants received a slow bolus intravenous injection of [18F]FEOBV with radioactivity doses ranging from 350-400 MBq. PET data acquisition started 180 minutes after injection, for a duration of 30 minutes, divided into 6 frames of 5 minutes each. All PET imaging sessions were supervised by a qualified nuclear medicine physician.

Participants reported a total of 53 adverse events of which 4 (7.5%) were related to the administration of [18F]FEOBV tracer. Of those 4 reports, one participant reported 2 events (mild dry mouth, mild strange sensations in the mouth) and a second participant reported 2 events (moderate nausea, moderate vomiting). Both recovered without treatment. No serious adverse events were reported.

### 2.4 PET and MRI data processing

SPM12 (http://fil.ion.ucl.ac.uk/spm/) for MATLAB was used for the following preprocessing steps. First, the six frames of the PET image were time-averaged to create a static PET image. The MRI scans were then linearly spatially normalized to the MNI152 asymmetrical template (<u>MNI152NLin2009cAsym template</u>) using SPM12’s unified segmentation algorithm (i.e., image registration, tissue classification, and bias correction). The same linear transformation matrices from the MRI normalization were applied to the PET images to align them with the MRI. Müller-Gärtner partial volume correction was applied to the PET data using the PETPVE toolbox, with an estimated point-spread function (PSF) of 2.4mm, to remove the partial volume effect on our PET images. The DARTEL module was used to estimate the nonlinear deformation field, and these were applied to the PET scan, further aligning the PET images to the MNI152 template space. To reduce noise, a Gaussian smoothing kernel with a full width at half-maximum (FWHM) of 6 mm was applied to the PET images. All preprocessing steps were conducted with appropriate quality control checks, ensuring data integrity and consistency throughout the process.

For all spatially normalized FEOBV PET images, the mean standard uptake value ratios (SUVR) were computed in pre-selected regions of interest (ROI) using a white matter mask as a reference region to normalize the PET images.^28^ Anatomical MNI-space Hammers atlas ^29^ was applied to the PET images to quantify regional differences in FEOBV binding in the anterior cingulate cortex (Matched MNI-space region: anterior cingulate), striatum (Matched MNI-space region: putamen and caudate), posterior cingulate cortex (Matched MNI-space region: posterior cingulate), primary sensorimotor cortex (Matched MNI-space region: precentral gyrus and postcentral gyrus), global cortex (frontal, temporal, occipital and parietal cortices), hippocampus (Matched MNI-space region: hippocampus), and parahippocampal gyrus (Matched MNI-space region: parahippocampal gyrus). The <u>Jülich Brain cytoarchitectonic atlas</u> was used to quantify the binding in the primary auditory cortex (Matched MNI-space region: TE1.0, TE1.1, TE1.2).

### 2.5 Outcome Measures

The endpoints relevant to the current paper include (1) baseline FEOBV binding in the *a priori* selected primary ROI (anterior cingulate cortex) and pre-specified exploratory ROIs (posterior cingulate cortex, primary auditory cortex, primary sensorimotor cortex, parietal lobe, frontal lobe, occipital lobe, temporal lobe, global cortex, hippocampus, parahippocampal gyrus, putamen, caudate, striatum), and (2) baseline cognitive performance using the executive composite z-score from the NIH EXAMINER battery.^30^

### 2.6 Statistical analysis

The distributions of the FEOBV SUVR for each ROI were tested for normality using the Shapiro-Wilk Test. To evaluate the association between FEOBV binding and age, we conducted Pearson correlations between baseline FEOBV SUVRs and age in years reported by the intent-to-treat (ITT) in the primary region of interest (i.e. anterior cingulate cortex) and exploratory ROIs. To evaluate the association between FEOBV binding and cognitive performance, we conducted Pearson correlations between baseline FEOBV SUVR in the primary region of interest (i.e. anterior cingulate cortex) and exploratory ROIs and the baseline cognitive composite z-score from the NIH EXAMINER battery.

No correction for multiple comparisons is made for exploratory analyses as specified in the published protocol. ^26^ All trending relationships (*p* < 0.10) are reported.

## 3. RESULTS

### 3.1 Participants

The flowchart of participants is depicted in Figure 1. Out of 113 individuals who were consented and screened, 20 were excluded for not meeting the criteria. Additionally, one participant was withdrawn by the site Principal Investigator before starting the assigned training program due to incidental findings on baseline imaging that precluded participation in the study. Thus, 92 participants completed the baseline neuropsychological assessments and neuroimaging sessions. Detailed information regarding the randomization procedures and intervention is available in the published protocol.^26^

**Figure 1.**
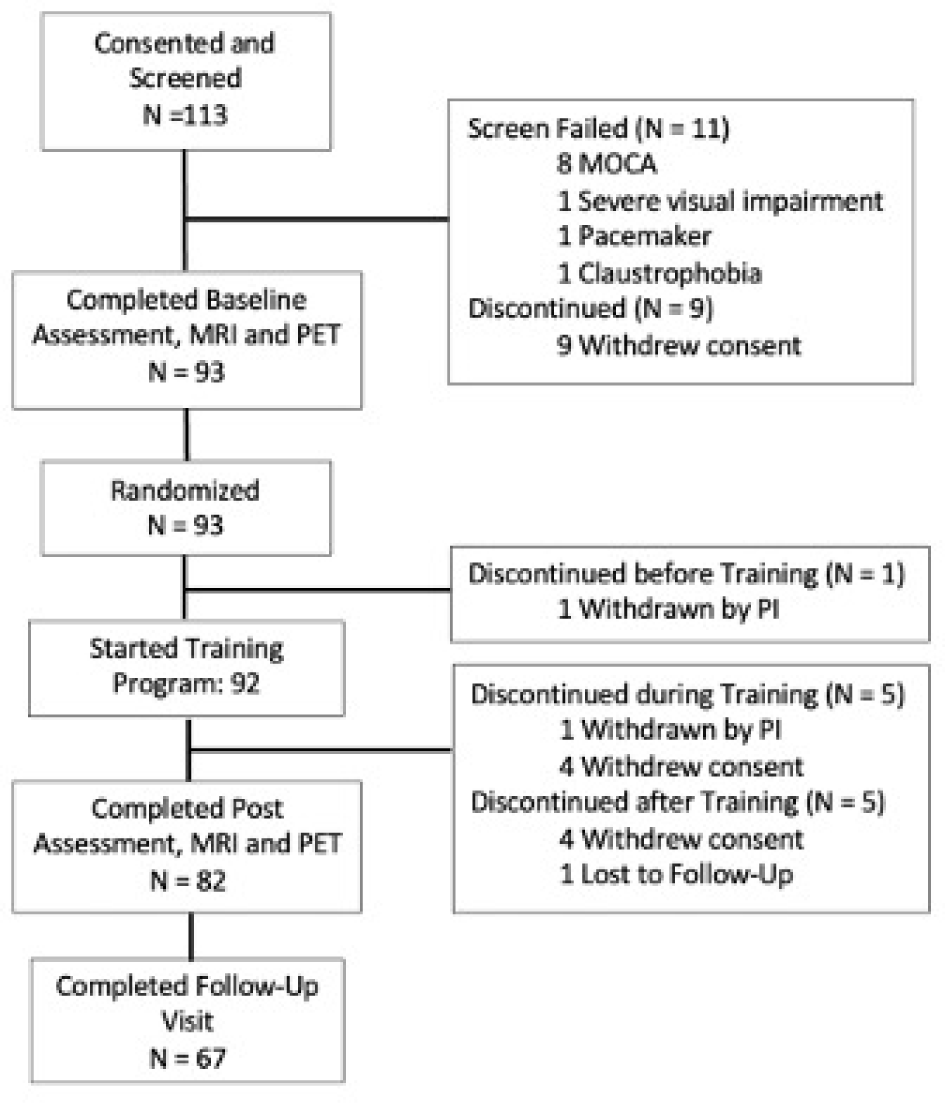
Flowchart for the INHANCE trial. Abbreviations: PI, Principal Investigator; MoCA, Montreal Cognitive Assessment; MRI, Magnetic resonance imaging; PET, Positron emission tomography.

### 3.2 Baseline characteristics of the intent-to-treat population

Demographic and neuropsychological characteristics for this study cohort (N= 92) are summarized in **Table 1**.

**Table 1.** Baseline characteristics of the intent-to-treat (ITT) population and all consented participants (N=113).

| **Participant's characteristics** | **Intent-to-treat (ITT) (N=92)** | **Consented (N=113)** |
| --- | --- | --- |
| Age, mean (SD), years | 71.9 (4.9) | 71.9 (5.0) |
| Education, mean (SD), years | 16.5 (3.4) | 16.7 (3.4) |
| Sex, n (%) |  |  |
| Female | 61 (66) | 71 (63) |
| Male | 31 (34) | 42 (37) |
| Race, n (%) |  |  |
| White | 88 (97) | 106 (93) |
| Asian | 2 (2) | 4 (4) |
| Black or African American | 1 (1) | 1 (1) |
| American Indian/Alaska Native | 0 (0) | 0 (0) |
| Native Hawaiian or Other Pacific Islander | 0 (0) | 0 (0) |
| More than one race | 0 (0) | 0 (0) |
| Unknown or not reported | 1 (1) | 2 (2) |
| Ethnicity, n (%) |  |  |
| Not Hispanic or Latino | 91 (99) | 110 (97) |
| Hispanic or Latino | 0 (0) | 2 (2) |
| Unknown or not reported | 1 (1) | 1 (0) |
| Native Language, n (%) |  |  |
| English | 33 (36) | 42 (37) |
| French | 50 (54) | 57 (50) |
| Other Language | 9 (10) | 14 (13) |
| Primary Language, n (%) |  |  |
| English | 43 (47) | 55 (50) |
| French | 48 (52) | 57 (50) |
| Other Language | 1 (1) | 1 (1) |
| Bilingual, n (%) | 12 (13%) | 18 (16%) |
| MoCA, total score (SD) | 26.2 (1.8) | Not applicable |
| GDS-SF, total score (SD) | 1.4 (1.9) | Not applicable |
| NIH EXAMINER, composite score (SD), raw | 0.44 (0.57) | Not applicable |

Abbreviations: ITT, intent-to-treat; SD, standard deviation; MoCA, Montreal Cognitive Assessment; GDS-SF, Geriatric Depression Scale - Short Form; NIH EXAMINER, National Institutes of Health The Executive Abilities: Measures and Instruments for Neurobehavioral Evaluation and Research,

### 3.3 Distribution of [18F]FEOBV PET signal

Brain FEOBV binding exhibited non-uniform distribution across various brain regions. The highest tracer binding was present in the striatum (putamen > caudate), followed by the hippocampus and cortical areas. The general distribution of [18F]FEOBV binding is described in **Table 2** and **Figure 2**.

**Figure 2.**
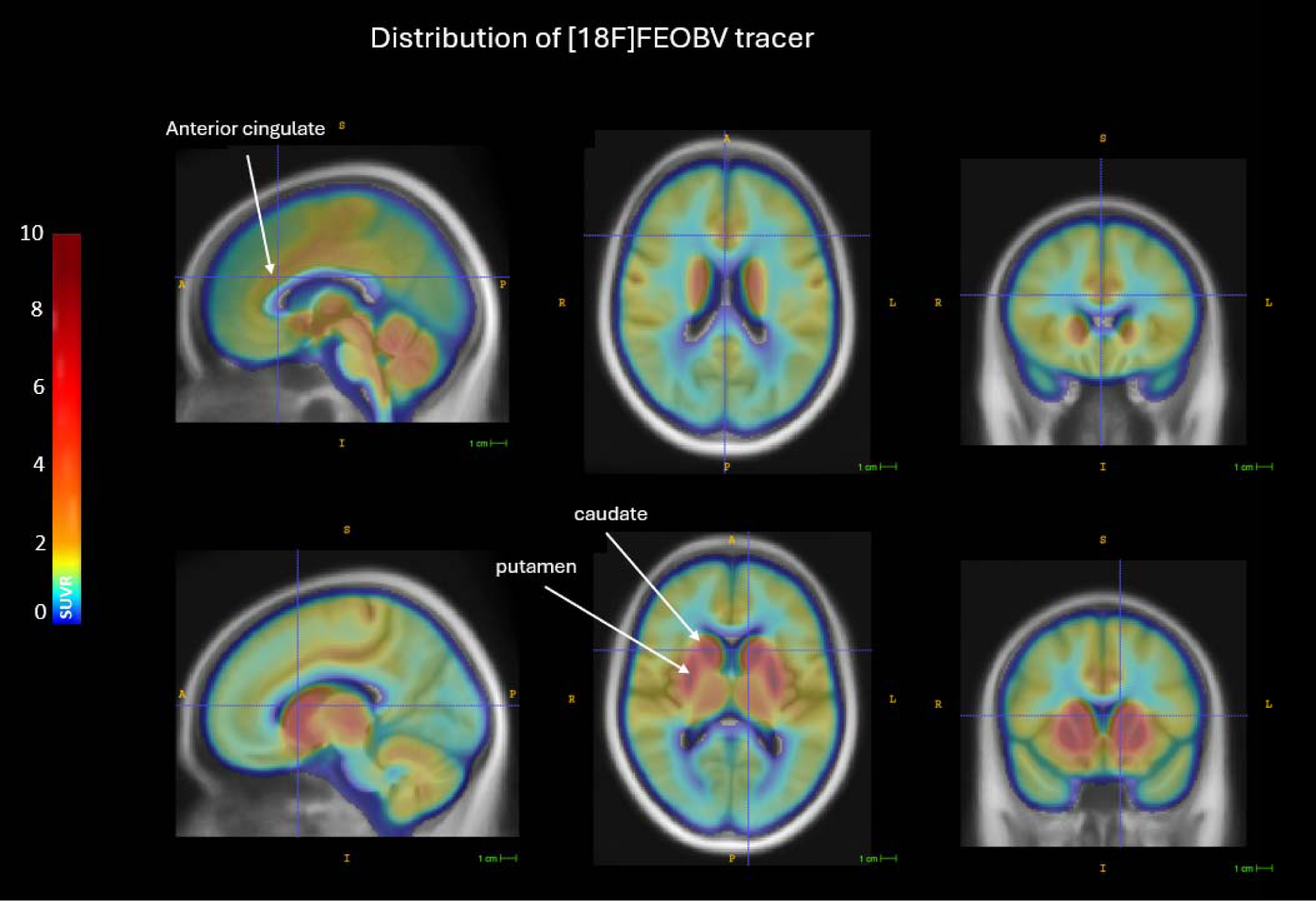
FEOBV binding topography in averaged brain slices of the intent-to-treat population (N=92). Abbreviations: A, Anterior; P, posterior; S, superior; I, inferior; R, right; L, left. SUVR: standard uptake value ratios.

**Table 2.** Pearson correlations between FEOBV binding, age and NIH EXAMINER across the pre-specified primary and exploratory ROIs for the ITT (N= 92) with bilaterally averaged FEOBV SUVRs and standard deviations (SD).

| Region of Interest (ROI) | Mean FEOBV<br>SUVR<br>(SD) | Correlations<br>with Age |  | Correlations with<br>NIH EXAMINER<br>executive composite |  |
| --- | --- | --- | --- | --- | --- |
|  |  | r | p | r | p |
| *Primary ROI* |  |  |  |  |  |
| Anterior cingulate cortex | 1.90 (0.19) | **-0.29** | **0.005** | **0.23** | **0.02** |
| *Exploratory ROIs* |  | r | p | r | p |
| Posterior cingulate cortex | 1.94 (0.20) | **-0.20** | **0.05** | 0.13 | 0.22 |
| Primary auditory cortex | 2.01 (0.30) | **-0.28** | **0.008** | 0.13 | 0.23 |
| Primary sensorimotor cortex | 1.73 (0.21) | -0.12 | 0.25 | *0.19* | *0.07* |
| Global cortex | 1.44 (0.14) | -0.19 | 0.07 | 0.16 | 0.13 |
| Parietal lobe | 1.46 (0.16) | -0.17 | 0.10 | 0.12 | 0.24 |
| Frontal lobe | 1.67 (0.17) | -0.11 | 0.29 | 0.07 | 0.48 |
| Occipital lobe | 1.20 (0.17) | -0.19 | 0.07 | *0.20* | *0.06* |
| Temporal lobe | 1.42 (0.17) | -0.17 | 0.10 | 0.14 | 0.17 |
| Hippocampus | 2.13 (0.29) | -0.13 | 0.21 | 0.14 | 0.18 |
| Parahippocampal gyrus | 1.39 (0.21) | -0.13 | 0.21 | 0.12 | 0.23 |
| Striatum | 6.09 (0.81) | **-0.26** | **0.01** | 0.12 | 0.27 |
| Putamen | 7.06 (0.98) | **-0.27** | **0.01** | 0.11 | 0.28 |
| Caudate | 5.11 (0.75) | **-0.21** | **0.05** | 0.11 | 0.30 |

Abbreviations: SUVR, standard uptake value ratios

### 3.4 Association between FEOBV binding and age for the intent-to-treat population

Baseline FEOBV SUVR was negatively associated with age in the anterior cingulate cortex, with lower uptake observed in older adults (r = -0.29, p = 0.005, **Figure 3**). This finding replicates previous research and confirms the validity of using FEOBV PET in this trial.^14^

**Figure 3.**
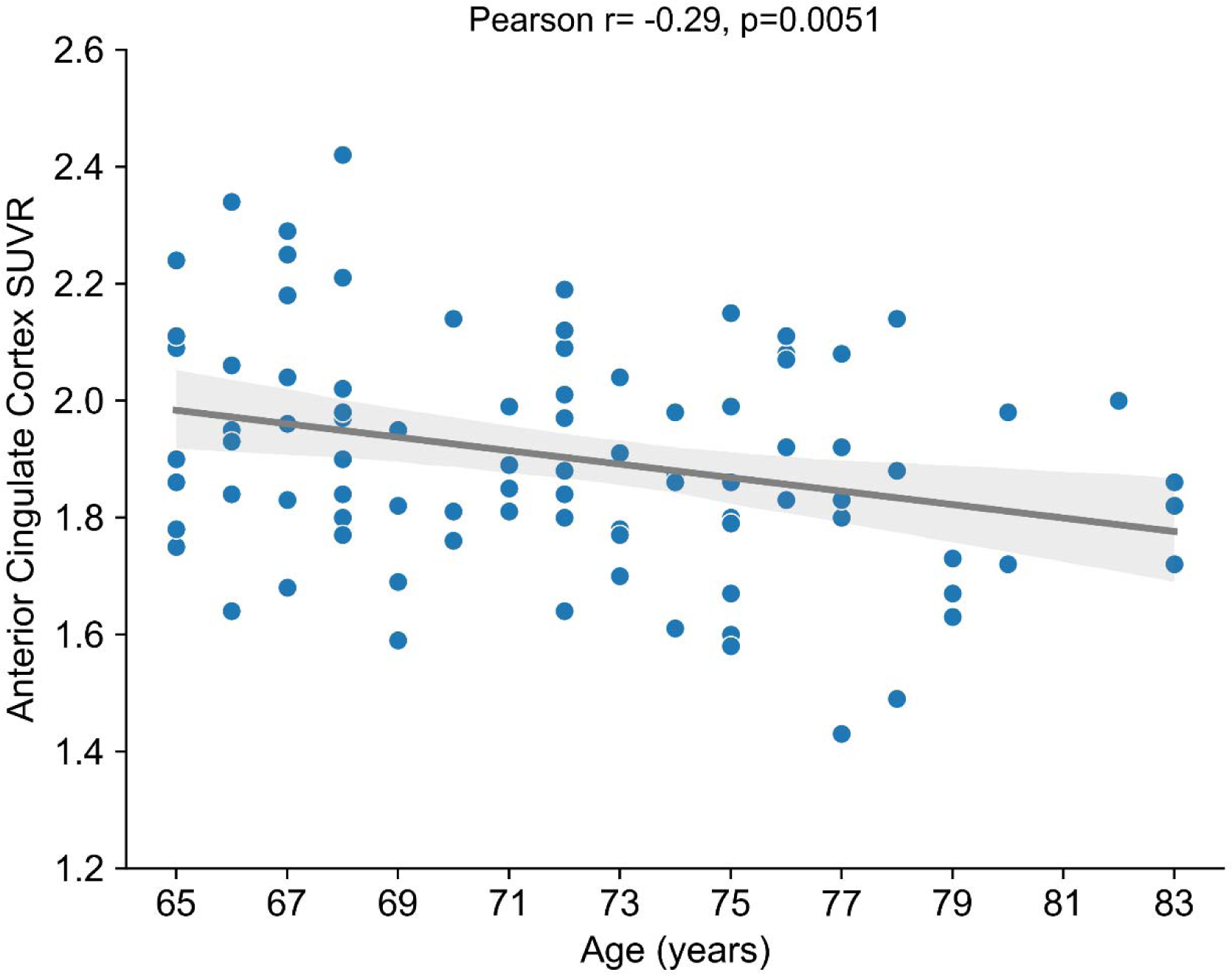
Pearson correlation between baseline FEOBV binding in the anterior cingulate cortex and age for the intent-to-treat population (N= 92). Each point represents an individual participant, with the regression line depicting the overall trend. Shaded regions around the regression lines represent 95% confidence intervals for the mean.

To further investigate predictors of FEOBV binding changes in the anterior cingulate cortex, a multiple linear regression analysis was conducted, including age, sex, and education as predictors. The overall model accounted for 8.6% of the variance in FEOBV binding (F(3, 87) = 2.73, p = 0.049, R= 0.29, R square= 0.086). Among the predictors, only age was a significant predictor of changes in FEOBV binding in the anterior cingulate cortex. The unstandardized coefficient indicates that for each additional year of age, FEOBV binding in the anterior cingulate cortex decreases by 0.011 units (B = -0.011, Beta = -0.28, t= -2.81, p = 0.006). With a baseline SUVR of 1.90, this corresponds to a 0.58% decrease per year and a cumulative decrease of 5.8% over a decade. In contrast, sex (B = 0.002, Beta = 0.004, t = 0.042, p = 0.96) and education (B = 0.001, Beta = 0.044, t = 0.42, p = 0.67) did not significantly affect FEOBV binding.

Baseline FEOBV SUVR values exhibited a significant negative association with age in several exploratory regions of interest (ROIs). Specifically, negative correlations were observed in the striatum (r = -0.26, p = 0.01), putamen (r = -0.27, p = 0.01), caudate (r = -0.21, p = 0.05), primary auditory cortex (r = -0.28, p = 0.008) and posterior cingulate cortex (r = -0.20, p = 0.05). No significant associations were found in the remaining ROIs. See **Table 2**.

### 3.5 Association between FEOBV binding and cognition for the intent-to-treat population

Baseline FEOBV SUVR in the anterior cingulate cortex exhibited a positive correlation with baseline NIH EXAMINER executive composite z-score (r = 0.23, p = 0.027, **Figure 4**). This indicates that reduced FEOBV binding in the anterior cingulate is associated with reduced cognitive performance. No significant associations were found across other exploratory ROIs. See **Table 2**.

**Figure 4.**
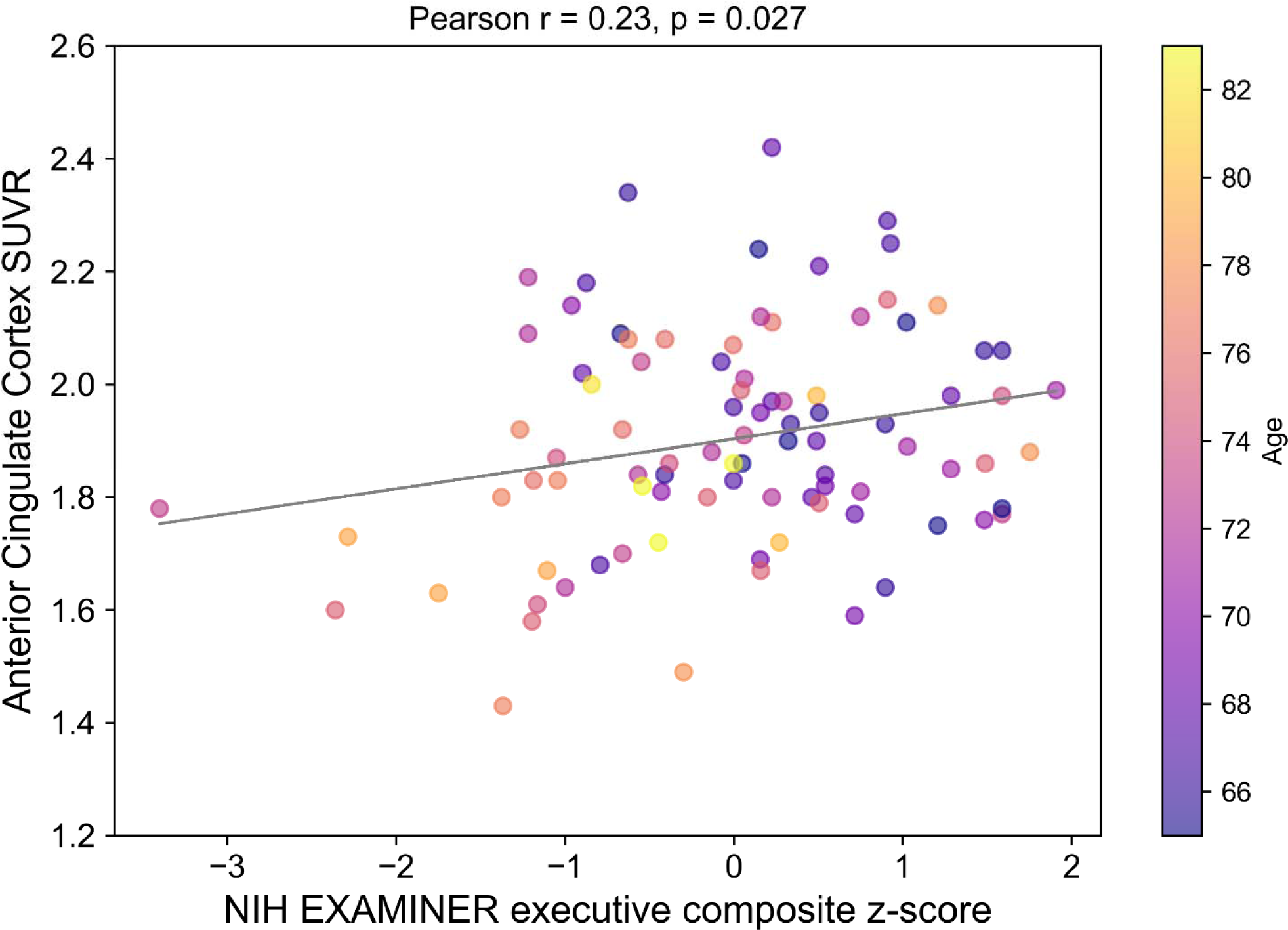
Pearson correlation between FEOBV binding in the anterior cingulate cortex and the NIH EXAMINER executive composite z-score for the intent-to-treat population (N=92) with darker observation points representing participants of relatively younger age. Abbreviations: SUVR, standard uptake value ratios.

## 4. DISCUSSION

In this study, we used [18F]FEOBV PET imaging to examine cholinergic nerve terminal distribution and its relationship with cognition in a large cohort of 92 neurocognitively intact older adults. We observed age-related reductions in FEOBV binding in the primary region of interest, the anterior cingulate cortex. Given the extensive projections of the basal forebrain cholinergic neurons throughout the cortex and subcortical regions, we conducted an exploratory analysis in additional ROIs. Significant age-related decreases in FEOBV binding were observed in the striatum (putamen and caudate), primary auditory cortex and posterior cingulate cortex. Additionally, FEOBV binding in the anterior cingulate cortex positively correlated with cognitive performance on the NIH EXAMINER executive composite score.

Current evidence suggests that [18F]FEOBV reliably assesses changes in the integrity of the brain’s acetylcholine system,^14,16,31,32^ with its binding distribution reflecting the known anatomical distribution of cholinergic terminals.^14,32–34^ In our study, the distribution of the [18F]FEOBV tracer was highest in the striatum, with greater binding observed in the putamen compared to the caudate, followed by the hippocampus and cortical regions. Overall, this heterogeneous cholinergic distribution is similar to previous histological post-mortem studies of brain cholinergic innervation in humans,^15,32,33^ as well as observed in rats ^34,35^ and non-human primates.^36^

We confirmed that aging is associated with decreased acetylcholine neurotransmission in the anterior cingulate cortex in healthy older adults, as indicated by reduced FEOBV binding in older individuals. Specifically, we observed approximately a 5.8% decline in FEOBV binding in the anterior cingulate cortex per decade. Similarly, Albin et al. recently used [18F]FEOBV PET imaging to investigate the topography of brain cholinergic innervation in 29 healthy subjects, with a mean age of 47 years (range 20–81 years). They reported age-related reductions in FEOBV binding, including approximately a 4% decrease per decade in the striatum and a 2.5% decrease per decade in both the primary sensorimotor cortex and anterior cingulate cortex.^15^ These results were later confirmed by a whole brain voxel-based analysis study in a larger sample with 42 healthy subjects (mean age 50 years, range 20–80 years).^31^ Unexpectedly, Okkels et al. found similar levels of regional FEOBV binding in a cohort of 20 neurocognitively intact older adults (mean age 74 years, range 64–86 years). These contrasting results may stem from variations in age ranges and the limited sample size used in Okkels’ study.^32^ Additionally, MRI morphometry studies have reported age-related reductions in basal forebrain volumes.^37^ Our results align with existing literature showing region-specific declines in FEOBV binding, suggesting changes in key basal forebrain cholinergic groups with aging. For instance, reductions in cholinergic terminals in the nucleus basalis of Meynert, which innervates the entire cerebral cortex.^33^ Nevertheless, this process is not uniform; we observed significant decreases in FEOBV binding in some cortical regions while others showed no age-related changes. Reductions in striatal cholinergic interneuron terminals, which provide the primary cholinergic input to the striatum,^33^ were also observed.

Furthermore, we investigated the relationship between FEOBV binding and NIH executive composite score. Notably, we observed a positive correlation between FEOBV binding in the anterior cingulate cortex and the NIH executive composite score in healthy older adults. To our knowledge, this is the first study to observe a positive association between FEOBV binding in the anterior cingulate cortex and cognitive performance in a large sample of neurocognitively intact older adults. Xia et al. reported positive correlations between global cortical FEOBV SUVR and cognitive composite scores for executive function (r = 0.70) and attention (r = 0.60) in a cohort of 18 individuals with MCI and cognitively intact older adults. For executive function, correlations were widespread across cortical and subcortical regions, while for attention, significant clusters were mainly in the anterior cingulate gyrus.^18^ Moreover, in a large sample of cognitively unimpaired Parkinson’s disease patients, positive associations were observed between global cortical FEOBV binding and cognitive performance across memory, executive functioning, and attention domains.^17^ Additionally, neurodegeneration in the nucleus basalis of Meynert preceded and predicted cortical degeneration and memory impairment in patients with Alzheimer’s disease.^12^

The mechanisms underlying age-related impairments in basal forebrain cholinergic neurotransmission are not fully understood. Possible biological factors may contribute to this decline. One key factor is the sensitivity of cholinergic neurons to disruptions in nerve growth factor signaling, which is crucial for their protection and maintenance. Such disruptions can lead to cellular atrophy and alterations in gene expression.^38^ Additionally, basal forebrain cholinergic neurons, characterized by their extensive axonal projections, are particularly susceptible to metabolic disturbances. The substantial energy requirements needed to maintain these widespread connections throughout life may underlie the vulnerability of the basal forebrain cholinergic system to neurodegenerative diseases.^39^ Given the critical role of these cholinergic neurons in important cognitive processes,^3^ the degenerative changes observed in cortical cholinergic terminals during normal aging likely contribute significantly to age-related cognitive decline.^40^ Indeed, augmenting acetylcholine levels through acetylcholinesterase inhibitors, has demonstrated cognitive enhancements in individuals with MCI and Alzheimer’s disease,^41,42^ underscoring the significance of acetylcholine in preserving the integrity of the brain and cognitive function.^43^ These findings collectively highlight the pivotal role of the cholinergic system in maintaining cognitive health and its potential as a target for interventions to mitigate age-related cognitive decline.

Limitations of this study include the use of a limited volume-of-interest analysis based on an a priori selection of regions with relatively age-related reductions in FEOBV binding reported in previous studies. A more spatially unbiased analysis would facilitate a more comprehensive assessment of the effects of aging on cholinergic neurotransmission.

## 5. CONCLUSION

In conclusion, we demonstrated that key cholinergic cell groups and their projections are particularly vulnerable to the effects of natural aging, with age-related changes in FEOBV distribution in the anterior cingulate cortex potentially playing a crucial role in cognitive decline. Our study adds to the literature by showing that [18F]FEOBV PET imaging is an effective marker for assessing changes in cholinergic neurotransmission, suggesting that maintaining an intact basal forebrain cholinergic system may provide cognitive resilience and protect against both age- and disease-related cognitive decline.

## Acknowledgements

We would like to thank the members of McGill’s Clinical Research and PET Imaging Units and the software engineers and quality assurance team at Posit Science.

## Author Contributions

Conceptualization: MA, EDS, TVV

Data curation: MA, ADFP, SJG

Formal analysis: ADFP, PJT, EDS, MA

Funding acquisition: MA, TVV

Investigation: SJG, LO

Methodology: MA, EDS, TVV, ADFP, PJT

Project administration: SJG, LO, PJT

Resources: MA, EDS, TVV

Software: MA, SJG

Validation: SJG

Supervision: MA, EDS, TVV

Visualization: ADFP, MA

Writing – original draft: ADFP

Writing – review and editing: ADFP, MA, EDS, TVV

## Data availability

Data available: Yes

Data types: Deidentified participant data (imaging, cognitive, behavioral)

When available: Two years after trial completion (on May 31, 2026), the INHANCE dataset will be made available to verified academic and industry researchers. Investigators can conduct both confirmatory and exploratory analyses using LORIS, a freely accessible, open-source data archive with provenance-sharing features. Access the dataset here: https://inhance.loris.ca/

